# Quantitative particle analysis of particulate matter release during orthodontic procedures: A pilot study

**DOI:** 10.1101/2020.09.09.20191270

**Authors:** Ahmed Riaz Din, Annika Hindocha, Tulsi Patel, Sanjana Sudarshan, Neil Cagney, Amine Koched, Jens-Dominik Mueller, Noha Seoudi, Claire Morgan, Shakeel Shahdad, Padhraig S. Fleming

## Abstract

**Introduction:** Transmission of SARS-Cov-2 through aerosol has been implicated particularly in the presence of highly concentrated aerosols in enclosed environments. It is accepted that aerosols are produced during a range of dental procedures, posing potential risks to both dental practitioners and patients. There has been little agreement concerning aerosol transmission associated with orthodontics and associated mitigation.

**Methods:** Orthodontic procedures were simulated in a closed side-surgery using a dental manikin on an acrylic model using composite-based adhesive. Adhesive removal representing debonding was undertaken using a 1:1 contra-angle handpiece (W&H Synea™ Vision WK-56 LT, Bürmoos, Austria) and fast hand-piece with variation in air and water flow. The removal of acid etch was also simulated with use of combined 3-in-1 air water syringe to remove etch. An Optical Particle Sizer (OPS 3330, TSI Inc. Minnesota, USA) and a portable Scanning Mobility Particle Sizer (NanoScan SMPS Nanoparticle Sizer 3910, TSI Inc Minnesota, USA) were both to assess particulate matter ranging from ‘very small’ (0.08 – 0.26 μm) to ‘large’ (2.7 – 10 μm) particles.

**Results:** Standard debonding procedure (involving air but no water) was associated with clear increase in the ‘very small’ and ‘small’ (0.26 – 0.9 μm) particles but only for a short period. Debonding procedures without air appeared to produce similar levels of aerosol to standard debonding. Debonding in association with water tended to produce large increase in aerosol levels, producing particles of all sizes throughout the experiment. The use of water and a fast hand-piece led to the most significant increase in particles. Combined use of the 3-in-1 air water syringe did not result in any detectable increase in the aerosol levels.

**Conclusions:** Particulate matter was released during orthodontic debonding, although the concentration and volume was markedly less than that associated with the use of a fast hand-piece. No increase in particulates was associated with prolonged use of a 3-in-1 air-water syringe. Particulate levels reduced to baseline levels over a short period (approx. 5 minutes). Further research within alternative, open environments and without air exchange systems is required.

## Introduction

Coronavirus disease 2019 (COVID-19) is caused by severe acute respiratory syndrome coronavirus 2 (SARS-CoV-2), wreaking havoc worldwide, with over 166 million confirmed cases and in excess of 875,000 reported deaths by September 5th, 2020.^1^ SARS-CoV-2 is transmitted primarily through direct or indirect contact with infected droplet. However, transmission of SARS-Cov-2 through aerosol has been implicated particularly in the presence of highly concentrated aerosols in enclosed environments.^2^

It is accepted that aerosols are produced during a range of dental procedures, posing potential risks to both dental practitioners and patients. This concern prompted decisions to cease elective dental procedures during the sustained human-to-human transmission phase of the COVID-19 pandemic, has led to implementing transmission-based infection prevention and control precautions including changes to personal protective equipment (PPE) requirements, and intensified international debate on the generation of aerosol within dentistry and orthodontics. Routine use of fallow periods to allow for settling of suspended aerosol has also variously been instituted following selected aerosol-generating procedures (AGPs) limiting the capacity for the provision of dental care. Notwithstanding this, there has been little consistency in the definition of an AGP, recommendation concerning PPE levels or the necessity for fallow periods following procedures.

The question of aerosol transmission has been contested within orthodontics with British national guidance implying that orthodontic treatment is essentially non-aerosol generating.^3^ Furthermore, the National Health Service defines a dental AGP as one “using high speed devices such as ultrasonic scalers and high-speed drills”^3^ with the use of slow-speed handpieces (SHP) not specifically mentioned. Orthodontic debonding and bonding procedures typically involve use of SHPs and conventional acid etch-based techniques, respectively, although alternatives do exist.^4^

Previous research into orthodontic-specific aerosol has focussed on procedures involving enamel clean-up associated with debonding of fixed appliances.^5–9^ Particulate generation has been attributed to the use of handpieces, burs, and water irrigants with a mass median aerodynamic diameter (MMAD) of up to 50μm,^6^ although particles with a MMAD of less than 0.75μm have also been noted.^9^ Smaller particles are of more concern, as they are expected to take longer to settle^10^ and are more likely to reach the terminal alveoli of the lungs.^7^ Particles with a MMAD of 0.5 to 10μm are considered to have the highest risk for transmitting infection.^11^ Respirable particulates can be produced during orthodontic debonding irrespective of whether a fast or SHP is used with or without water irrigation.^9^ However, fast handpieces (FHPs) used with water irrigation are considered likely to induce the most aerosol production.^8^ It has also been suggested that pre-procedural mouth-rinses may offer no measurable benefit.^6^ The make-up of released particles incorporating composite resin, filler and metallic material has also been illustrated.^5,12,13^

In previous studies, qualitative sampling of particle emission allowing assessment of potential lung penetration and the collection of respirable particulate fractions has been undertaken with a Marple Cascade Impactor (Thermo Fisher Scientific^TM^, Franklin USA).^7–9^ This monitor has also allowed for scanning electron microscopy and energy dispersive x-ray spectroscopic (EDX) analysis.^7^ Other cascade impactors have also been used such as the Six-Stage Viable Anderson Cascade Impactor (Thermo Fisher Scientific^TM^, Franklin USA), which has been used to represent six stages of the respiratory tract.^6^ With regards to quantitative measurements, a Personal DataRAM (pDR) 1200 Real Time Monitor (Thermo Fisher Scientific^TM^, Franklin USA) which is a form of photometric monitor, has been used to measure the concentration of respirable particles.^7,8^ Markers such as fluorescence and citric acid used in water lines and the spread of splatter have been used as surrogate measures of dental aerosol release.^14–16^ Alternatives have included attempts to identify environmental contamination using microbiological markers by cultured air sampling techniques.^14,17^ However, these approaches are designed to identify the characteristics and behaviour of aerosol produced in a dental operatory. Fluid dynamic methods involving more sensitive particle analysis offer the potential to identify the characteristics and behaviour of an aerosol including the size, spread and longevity of particles following dental procedures.^12,18,19^ Few studies in dentistry have used particle analysis for assessment of aerosol.^13,18^

Previous research has failed to elucidate the quantity and nature of particles produced following orthodontic procedures, and the duration required for particulates to reduce to baseline levels. This information would be helpful in determining the potential risk of SARS— CoV-2 from aerosol production during orthodontic procedures including hand piece-based debonding and the use of 3-in-1 air water syringes. The aim of this pilot study was therefore to identify whether aerosol is produced during orthodontic procedures and, if so, to estimate the duration of this effect. Secondary aims were to clarify the intensity of the aerosol produced including the range of particle sizes, and to explore whether the use of water during debonding enhances or reduces emitted aerosol.

## Methodology

Orthodontic treatment procedures were simulated in a closed side surgery in a dental hospital setting (Institute of Dentistry, Barts and The London School of Medicine and Dentistry) using a dental manikin. The dental hospital has an air exchange system (with six air changes per hour) which was left functioning throughout the procedures.

A 1mm layer of composite based adhesive (3M^TM^ Transbond^TM^ XT Light Cure Paste Adhesive, Minnesota USA) was light-cured on the buccal surface of all upper and lower acrylic teeth including second molars (28 teeth in total). Adhesive was placed using orthodontic attachments with the base coated in petroleum jelly with 300g of pressure applied for 5 seconds. Excess was removed and the adhesive light-cured for 20 seconds per tooth with an light emitting diode (LED) curing light. A permanent black marker pen was used to identify the location of the adhesive and demarcate the surface of the manikin (Figure 1).

**Figure 1.**
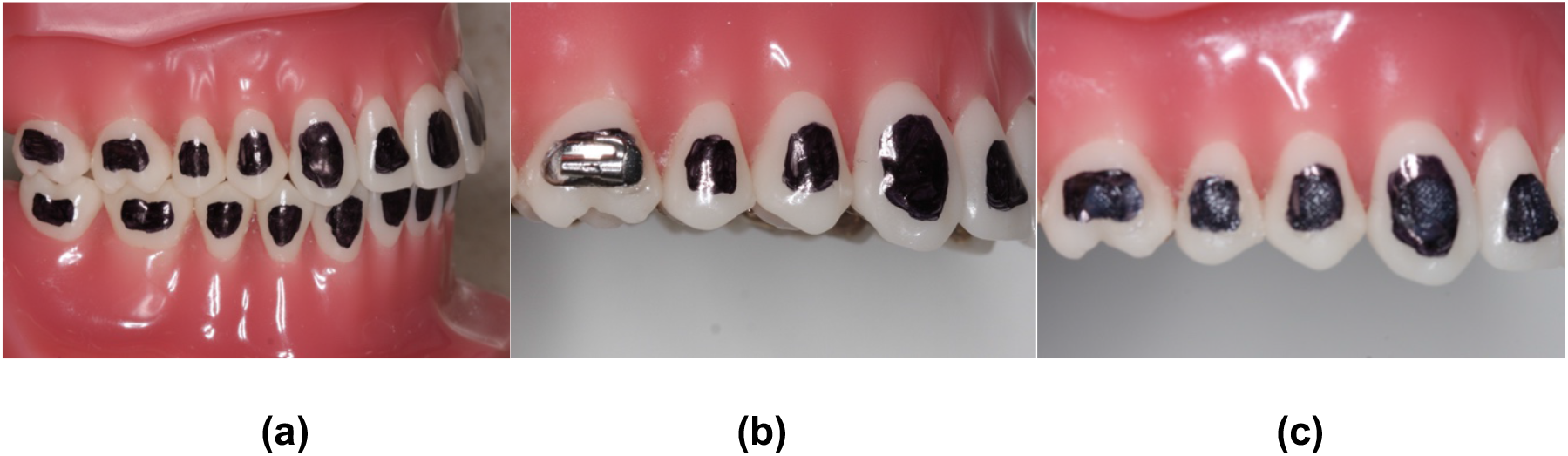
Experimental model to simulate debonding with the location of adhesive demarcated (a). Adhesive was placed on the tooth surface with pressure to expel the excess adhesive using molar tubes and brackets as appropriate covered in a petroleum jelly layer (b). An even layer (1mm) of composite-based adhesive remained on each tooth surface (c).

Debond procedures were simulated with adhesive removal using a 1:1 contra-angle handpiece (W&H Synea^TM^ Vision WK-56 LT, Bürmoos, Austria); handpieces were operated at maximum speed with and without the use of air pressure and water flow. Experiments were also undertaken using 37% phosphoric acid etch placed on all 28 teeth with use of combined 3-in-1 air water syringe to remove etch. A two-bracket repair was also simulated with adhesive removal from two teeth (premolar and incisor). All experiments were undertaken using four-handed dentistry technique using high volume suction (HVS). The operator and assistant only were present in the room during the pre-procedural period and left on completion of the procedure. Both operator and assistant wore fluid resistant surgical masks (FRSMs) during the procedures. All procedures were undertaken by the same operator (PSF) in the same closed surgery. Procedures were repeated on separate days over a period of 3 weeks to ensure that variation in external factors pertaining to air exchange and air-conditioning systems were accounted for.

An optical particle scanner (OPS 3330, TSI Inc. Minnesota, USA) and a spectrometer particle scanner (NanoScan SMPS 3910, TSI Inc. Minnesota, USA; Figure 2) were used to assess particulate matter. The sampling inlets were located at a fixed position at 7 o’clock and located 8cm away from the maxillary left central incisor (Aerosol Instrument Manager AIM^®^v10.3.1.0 and NanoScan Manager^®^ v1.0.0.19, TSI Incorporated, Minnesota, USA) with a sampling rate of one minute. Measurement commenced 10 minutes prior to the procedure to assess the baseline concentration of particulate matter and continued for 30 minutes following the procedure. Using both counters it was possible to measure particles in 26 bins, ranging from 10nm to 10μm in diameter. As a single SAR-CoV-2 viron is approximately 80 to 100 nm in diameter,^20^ information of particles smaller than 80nm was deemed to be irrelevant to virus transmission and was discarded. In order to reduce the remaining data, the particle counts were cokmbined into four categories: ‘very small’ (0.08 – 0.26 μm), ‘small’ (0.26 – 0.9 μm), ‘medium’ (0.902 – 2.7 μm) and ‘large’ (2.7 – 10 μm).

**Figure 2.**
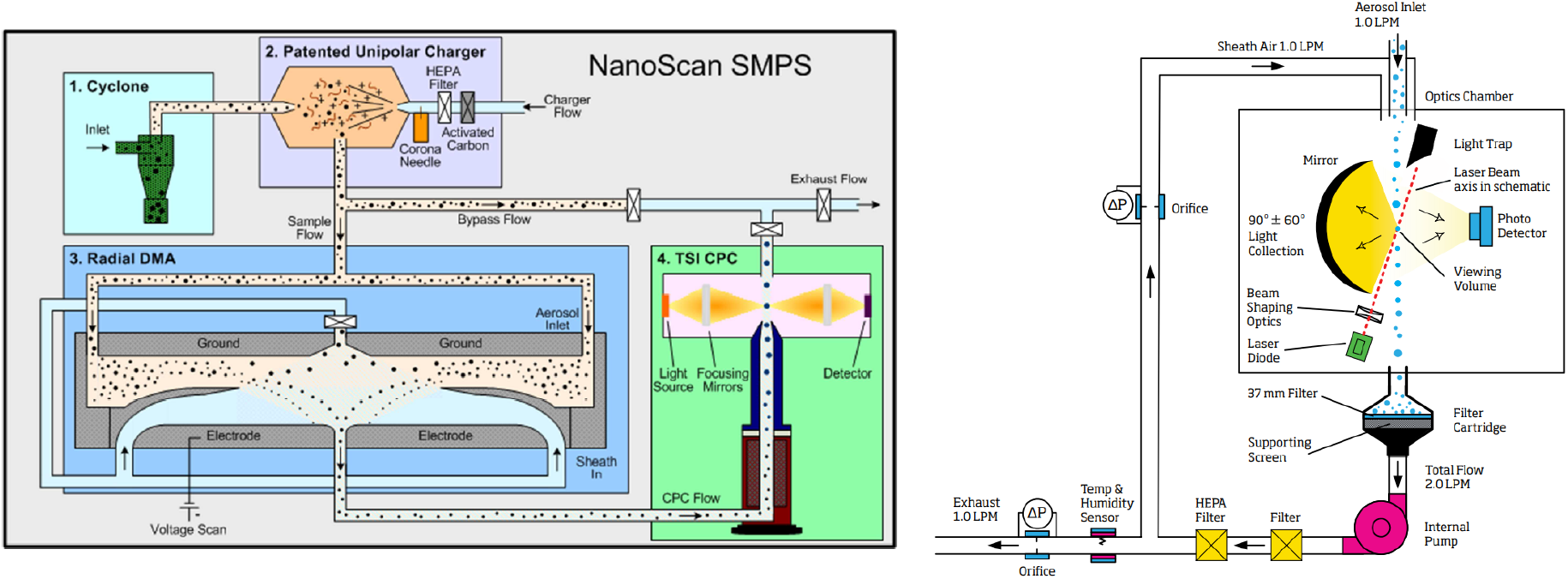
Working principle of the instruments a) NanoScan 3910; b) OPS 3330

Tests were first inspected to ensure that aerosol levels were stable during the initial fallow period, and cases in which large fluctuations were observed were disregarded. To quantify any changes in the particle concentration, the median of the concentration of particles in each size range during each procedure was calculated and was compared to the corresponding value in the initial 10-minute fallow period prior to each procedure. The Mann-Whitney U-test was applied to estimate the probability that the median of the measurements acquired during the procedure was different to that of the fallow period (i.e. that the concentration of aerosols had increased). In this context, a high P-value indicates a high likelihood that the procedure has led to a change in the median aerosol concentration. This statistical test was not applied to the 3-in-1 air water syringe used with etch or the breakage repair, as these procedures took less than 1 minute and therefore represented a single data point.

## Results

Most procedures ranged in duration from 4 to 11 minutes for debonding using either air, no air, water or FHP (Table 1), with the exception of washing of enamel with 3-in-1 air water syringe and the repair of two teeth, which both took less than 60 seconds. Data showing the percentage change in the median concentration of aerosols during each procedure (relative to the initial fallow time) and the probability that this change is statistically significant (based on the Mann-Whitney U-test) are presented in Tables 1 and 2, respectively. Most procedures led to a statistically significant increase in aerosol concentrations in most size ranges (Tables 1 and 2).

**Table 1.** Percentage change in the median concentration of particles in various size ranges during the procedures, with respect to the baseline levels (measured in the 10 minute period prior to the start of the procedure). For the final two procedures (repair and the use of 3-in-1 air water syringe used to remove etch), the values show the percentage change in the single data point recorded during the procedure, relative to the baseline.

| Procedure | Number | Duration (mins) | Very Small | Small | Medium | Large |
| --- | --- | --- | --- | --- | --- | --- |
| | | | 0.08 – 2.6 $\mu\text{m}$ | 0.26 – 0.9 $\mu\text{m}$ | 0.9 – 2.6 $\mu\text{m}$ | 2.6 – 10 $\mu\text{m}$ |
| **Standard Debond** | 1 | 11:00 | 104.7 | 111.7 | 97.3 | 136.7 |
|  | 2 | 11:00 | 104.4 | 119.5 | 82.0 | 78.7 |
|  | 3 | 4:00 | 144.2 | 246.0 | 155.1 | 86.3 |
|  | **Average** |  | **117.8** | **159.1** | **111.5** | **100.6** |
| **Debond without Air** | 1 | 4:10 | 152.9 | 344.1 | 149.4 | 86.1 |
|  | 2 | 5:00 | 171.8 | 140.4 | 225.5 | 367.8 |
|  | 3 | 4:00 | 328.3 | 410.2 | 3040.3 | 5195.5 |
|  | **Average** |  | **217.7** | **298.2** | **1138.4** | **1883.1** |
| **Debond with Water** | 1 | 9:00 | 96.0 | 133.2 | 161.1 | 89.4 |
|  | 2 | 3:50 | 100.8 | 246.7 | 1523.9 | 6831.7 |
|  | **Average** |  | **98.4** | **190.0** | **842.5** | **3460.5** |
| **Debond with FHP** | 1 | 11:00 | 135.2 | 84.8 | 150.0 | 430.1 |
|  | 2 | 11:00 | 96.9 | 146.7 | 294.4 | 179.8 |
|  | 3 | 11:00 | 89.1 | 140.9 | 401.5 | 168.2 |
|  | **Average** |  | **107.1** | **124.1** | **282.0** | **259.4** |
| **Repair**<br>**(Standard**<br>**Debond)** | 1 | 0:30 | **415.9** | **328.8** | **668.5** | **103.4** |
| **3-in-1 air**<br>**water**<br>**syringe** | 1 | 0:30 | **81.2** | **94.3** | **157.5** | **110.7** |

**Table 2.** Probability that there has been a change in the median particle concentration measured during a procedure relative to baseline. Probabilities were calculated using the Mann-Whitney U-test, with values close to 1 indicating a high probability that the dental procedure led to an increase in aerosols within that size range.

| **Procedure** | **Number** | **Duration**<br>**(mins)** | **Very Small** | **Small** | **Medium** | **Large** |
| --- | --- | --- | --- | --- | --- | --- |
|  |  |  | **0.08 – 2.6 µm** | **0.26 – 0.9 µm** | **0.9 – 2.6 µm** | **2.6 – 10 µm** |
| **Standard**<br>**Debond** | 1 | 11:00 | 0.94 | 0.99 | 0.12 | 0.55 |
|  | 2 | 11:00 | 0.66 | 0.9 | 0.06 | 0.07 |
|  | 3 | 4:00 | 0.83 | 0.67 | 0.89 | 0.87 |
|  | **Average** |  | **0.81** | **0.85** | **0.36** | **0.50** |
| **Debond**<br>**without Air** | 1 | 4:10 | 0.72 | 0.97 | 0.98 | 0.48 |
|  | 2 | 5:00 | 1 | 1 | 1 | 1 |
|  | 3 | 4:00 | 0.97 | 1 | 1 | 1 |
|  | **Average** |  | **0.90** | **0.99** | **0.99** | **0.83** |
| **Debond**<br>**with Water** | 1 | 9:00 | 0.18 | 1 | 1 | 0.4 |
|  | 2 | 3:50 | 0.89 | 0.98 | 1 | 1 |
|  | **Average** |  | **0.69** | **0.66** | **0.67** | **0.54** |
| **Debond**<br>**with FHP** | 1 | 20:00 | 0.99 | 0.23 | 0.98 | 1 |
|  | 2 | 20:00 | 0.73 | 1 | 1 | 0.9 |
|  | 3 | 20:00 | 0.1 | 1 | 1 | 0.97 |
|  | **Average** |  | **0.61** | **0.74** | **0.99** | **0.96** |

The use of hand-piece based debonding resulted in an increase in particulate matter (Figure 3(a) and Table 1). A standard debonding procedure (involving air but no water) was associated with a clear increase in the ‘very small’ and ‘small’ particles, but only for a short period with no significant increase for other sizes. Debonding procedures without air appeared to produce similar levels of aerosol to standard debonding. A sharp increase in particles at the start of one procedure with air was observed most likely related to a misalignment of the handpiece; this appears to be an outlier. However, all three repeats show a consistent increase in the concentration of ‘small’ and ‘medium’ particles (Figure 3(b,ii), Figure 3(b,iii) and Table 2). Debonding with water tended to result in large peaks, producing particles of all sizes throughout the experiment. The aerosol increase during debonding with water appeared to be of a higher magnitude than for standard debonding procedures due to the production of both solid particles and droplets of water.

**Figure 3.**
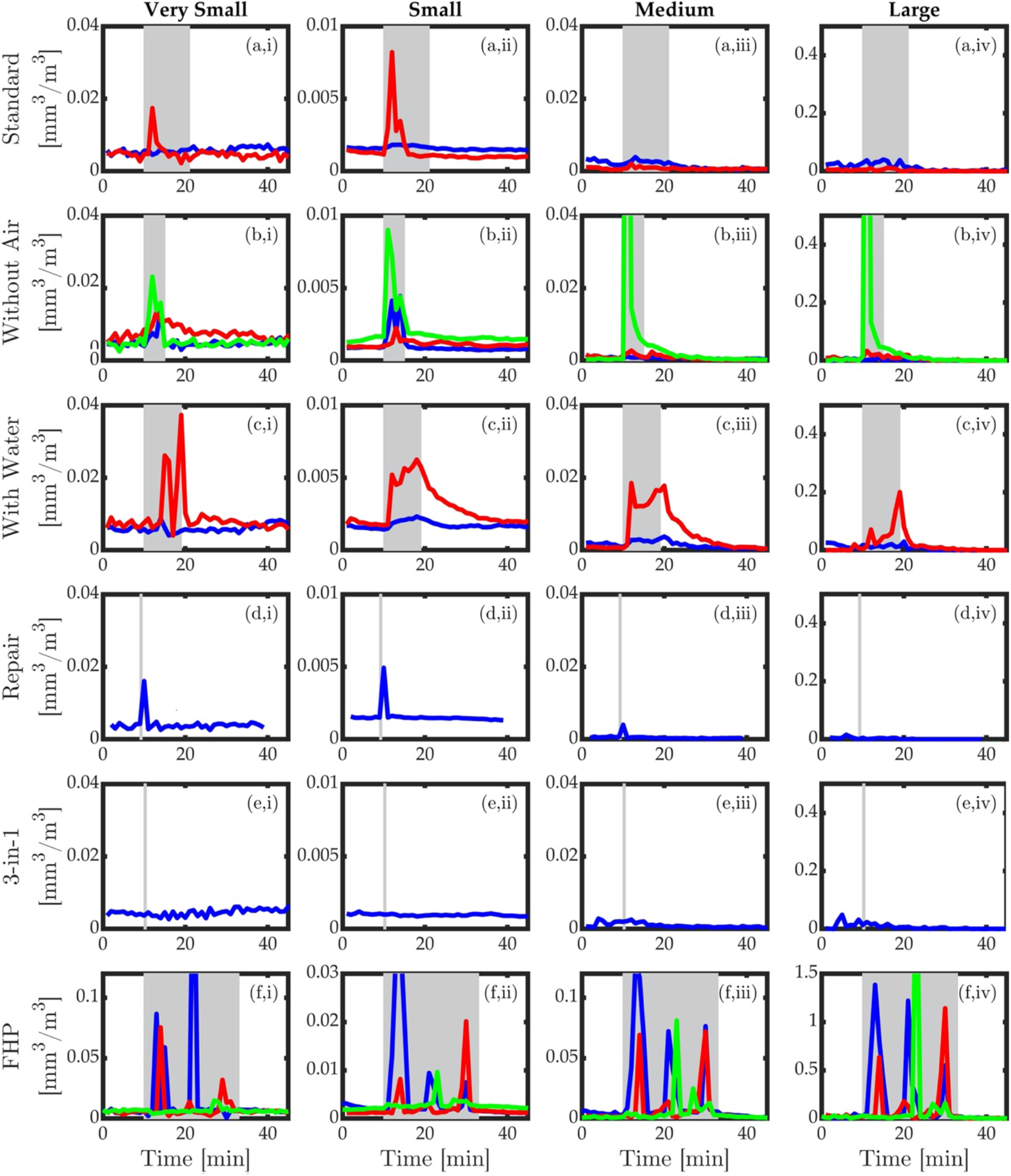
Variation in the measured concentrations of aerosols during each experiment. The rows correspond to the different procedures: standard debond (a), debond without air (b), debond with water (c), repair of two teeth using standard debond approach (with air only) (d), and 3-in-1 air water syringe (e) and debond using the fast-hand piece (f). The columns show the concentrations for various size ranges of aerosols; ‘very small’ (i), ‘small’ (ii), ‘medium’ (iii) and large ‘(iv)’. The grey shaded region indicate the time during which the procedure took place, and the colours indicate the different repeated measurements, as shown in Table 2; 1 (blue), 2 (red) and 3 (green).

*Note that the y-axis scale is different for the FHP procedures (Row f).

The use of a FHP with water led to the most significant increase in particles. However, in this case the increase was associated with short spikes that were very large, rather than a continuous increase, as was seen in previous cases. An additional test was performed to assess the aerosol produced during a repair procedure, in which the standard debond (with air but no water) was applied to only two teeth (Figure 3(d)). As in the case of the complete debond (Figure 3(a)), this was associated with an increase in aerosol concentration in the ‘very small’ and ‘small’ size ranges, although the relatively short duration meant the total increase in concentration is much smaller. Finally, the combined use of 3-in-1 air water syringe to remove etch did not result in any detectable increase in the aerosol levels.

## Discussion

The aerosol dilemma has been thrown into sharp focus since the onset of the Covid-19 pandemic with binary discussion around the presence or otherwise of aerosol associated with various dental procedures.^3^ There has also been consideration of the potential risk attached to various procedures. This risk-based approach appears to be relevant in relation to orthodontics, in particular. Specifically, the majority of orthodontic procedures may lead to splatter while not producing aerosol. However, on the basis of the present study, it is apparent that SHP-based debonding procedures do generate aerosol, although the magnitude of aerosol release is markedly lower than that emitted by FHP use.

The nature of the aerosol released during debonding has not been fully quantified in this analysis. However, given that no salivary substitute was used, it is likely to constitute dry debris in the absence of the use of water. The viability of envelope viruses such as SARSCoV-2 is known to be reduced when virons are not in a liquid environment^21^; however, in a clinical setting, viral particles are likely to reside in or be mixed with saliva, increasing the potential for transmission. Furthermore, a significant volume of ‘small’ and ‘very small’ particles were measured during orthodontic treatment procedures, in keeping with previous research.^8^ These are within inhalable and respirable fractions and it is believed that such droplets can conceivably lead to transmission of SARS-CoV-2.^22^

The release of fine particulate matter during full debonding procedures was observed consistently. Debonding under irrigation was also evaluated as it was hypothesized that adjunctive use of water might increase the mass and volume of any generated aerosol making it more amenable to removal with high-volume suction. A beneficial effect of irrigation, however, was not observed. Moreover, the performance of debonding with reduced air pressure on the SHP did not eliminate aerosol release. It therefore appears that debonding procedures involving the use of SHPs will inevitably lead to some aerosol release. Notwithstanding this, the concentration and mass of released particles using the SHP for debonding was markedly lower than that observed with the use of FHPs. Moreover, particulate levels reduced to background levels within a period of 5 to 10 minutes. It is important to note that the air filtration system within the environment assessed is likely to have facilitated this rapid dispersal. As such, given the time necessary to complete an orthodontic debond including scanning or impression taking, a post-procedural surgery fallow period is unlikely to be necessary. This may not, however, be true of all closed surgical environments where air filtration facilities are lacking.

The findings from the present study are consistent with previous research identifying aerosol and splatter, which may be a possible route for microbial transmission within a clinical environment.^11,18^ Two primary clinical considerations exist: how to protect the treating clinician and assistant from the aerosol released, and whether aerosol emitted poses a risk to subsequent patients. The magnitude of the risk associated with orthodontic debonding is difficult to quantify; notwithstanding, given the low levels of aerosol produced with the SHP relative to FHP use, it would be reasonable to infer a commensurately lower risk profile. However, the routine use of enhanced PPE and in particular masks would be sensible whether the FHP or SHP is used for adhesive removal during debonding. FRSMs have been shown to have limited value in relation to dust resistance with a quoted 96% reduction pertaining to fluid resistance, in isolation.^8^ Furthermore, Type I and Type IR face masks have a bacterial filtration efficiency (BFE) of 95%, whereas Type II and Type IIR face masks have a slightly higher BFE of 98%.^23^ These medical masks are tested in the direction of exhalation (inside to outside) and take into account the efficiency of bacterial filtration. Surgical masks of this type protect from droplet transmission of the infection but will not offer adequate protection against close range aerosol transmission for viruses such as coronavirus.^23^ As such, further refinement and guidance in relation to the appropriate mask protection is required.

In terms of the risk attached to orthodontic debonding, it is important to note that differences exist between removal of superficial composite material and operative dentistry. The latter may involve excavation of infected dentine and enamel, and pooled microbial communities; however, debonding is based on removal of a surface composite layer where infected dental tissue is not encountered. The associated risk on the production of harmful bioaerosol is, however, uncertain with SARS-CoV-2 receptors present in salivary glands and the pharynx with salivary and pharyngeal secretion, therefore, potentially infected. It is worth noting that excess adhesive flash surrounding orthodontic attachments may be exposed to the oral environment for some time. This can be limited by limiting adhesive flash; although this has failed to reduce the production of particulates in the inhalable, respirable or thoracic fractions has been demonstrated.^7^

The present study has highlighted the potential for relatively short-lived and low concentration of aerosol release during orthodontic debonding. While quantification of risk is fraught, it would be reasonable to assume that this is likely to be limited given the lower particulate volumes and concentration for orthodontic debond using SHPs relative to procedures involving FHPs. In certain jurisdictions, decisions were made during the first wave of the COVID-19 pandemic to cease elective appointments and indeed orthodontic emergencies presumably on the premise of the ‘precautionary principle’, to mitigate against viral spread given the non-zero probability of risk. It is accepted, however, that orthodontics involves an ongoing course of treatment with recognised risks associated with noncompliance and unsupervised care, including unwanted tooth movement and iatrogenic damage.^24^ As such, while remote monitoring offers some promise in mitigating potential associated problems, the risks associated with in-person attendance should be balanced with the undeniable need for progression of treatment and physical attendance on a regular basis during active courses of treatment.^25^

A significant limitation of the present study was that the particulate model did not measure viral load generated from these procedures. Dental aerosols are distinct from respiratory aerosols as they produce their own external aerosol with the potential for infectivity residing in the patient’s mucus, nasal and oral secretions. The air and water jets produced by FHPs and SHPs in the oral cavity may become contaminated; the associated microbial concentration and risk of infectivity has not yet been elucidated fully. While quantification of the precise microbial contamination and infectious dose of the particles assessed was not possible, one can estimate the risk based on the known ability of the smaller particles to persist longer in the air and penetrate deeper in the respiratory tract. In general, smaller particles are more likely to be airborne; as such, the risk of airborne spread was considered.^26^

## Conclusions

Based on this simulated study undertaken within a closed surgery, particulate matter is released during orthodontic debonding. The concentration and volume of matter produced using the SHP for adhesive removal during debonding is markedly less than that associated with FHP use. No increase in particulates was associated with prolonged use of a 3-in-1 air water syringe. Particulate levels reduced to baseline levels over a short period (approximately 5 minutes) in this environment. Therefore, in the current SARS-CoV-2 risk environment we recommend that orthodontic debonding be carried out using the SHP rather than the FHP, and without the use of water as an irrigant, to mitigate the risk of infection to staff and patients. Whilst it is difficult to extrapolate to other air environments, in this setting, the fallow periods between patients can be reduced to approx. 5 minutes and use of the 3-in-1 air water syringe considered to be a low-risk dental procedure.

## Data Availability

All data is available for review

